# The Effects of Mindfulness Meditation on Burnout in Clinical Genetic Counselors: A Three-Arm Randomized Controlled Trial

**DOI:** 10.64898/2026.01.14.26344130

**Authors:** Colleen Caleshu, MaryAnn Campion, Jehannine (J9) Austin, Philippe Goldin, Julia Silver, Aad Tibben

**Affiliations:** Genome Medical, South San Francisco, CA, USA; Centre for Human and Clinical Genetics, Leiden University Medical Centre (LUMC), Leiden, The Netherlands; Stanford University School of Medicine, Department of Genetics, Stanford, CA, USA; University of British Columbia, Departments of Medical Genetics and Psychiatry, Vancouver, BC, Canada; Betty Irene Moore School of Nursing, University of California, Davis; University of California San Francisco, Prenatal Diagnostic Center, San Francisco, CA, USA

## Abstract

Burnout is common among genetic counselors (GCs). Clinician burnout has been found to adversely affect individual well-being, patient care, and likelihood of staying in a role. Both individual and systems solutions are needed to address clinician burnout. Mindfulness meditation (MM) is one individual-level solution that has shown promise for reducing burnout in other clinicians but has not been studied in GCs. We conducted a decentralized, parallel, three-arm randomized controlled trial comparing MM to a novel active control meditation (ACM) and a no-meditation control (NMC), with 1:1:1 randomization. Participants were clinical GCs in the US. MM and ACM participants were asked to do 10 minutes of daily app-based meditation for 8 weeks. ACM was designed to control for non-specific aspects of MM by mimicking MM length and structure without including mindfulness techniques. The primary outcome, burnout, was assessed using the Professional Fulfillment Index. Secondary outcomes included other indicators of professional well-being, such as stress and professional fulfillment. Outcomes were assessed via an intention-to-treat approach, with multiple imputation for missing outcome data. Outcome analyses controlled for baseline trait mindfulness. 397 participants (mean age 33 years; 97.7% female, 94.2% White) were randomized, and 76% completed post-intervention outcome measures. There was no difference in burnout reduction between MM and ACM groups (p = 0.44). However, multiple measures suggest that ACM did not perform well as an inert active meditation control, thus the primary hypothesis could not be effectively tested. In pre-planned secondary analyses, MM reduced burnout (Cohen’s d = −0.84, p < 0.001) compared to NMC, a passive control. Similar results were seen for stress. These findings suggest MM may be beneficial for GC professional well-being; however, further research on MM for GCs is needed with more diverse study samples and better active controls.

## INTRODUCTION

Burnout is common among genetic counselors (GCs), with a point prevalence over 50% (Bernhardt et al. 2009; Udipi et al. 2008; Lee et al. 2015; Allsbrook et al. 2016; Johnstone et al. 2016; Caleshu et al. 2022). Burnout is characterized by emotional exhaustion, depersonalization, and reduced sense of personal accomplishment (Maslach 2001). Beyond its impact on individual well-being, burnout is a significant factor in clinicians leaving direct patient care (Williams et al. 2001; Hayes et al. 2006; National Academies of Sciences, Engineering, and Medicine 2019). GCs consistently report burnout as the primary reason they consider leaving the field (National Society of Genetic Counselors 2023). Furthermore, clinician burnout has been associated with decreased quality of patient care and increased medical errors across healthcare settings (Shanafelt et al. 2010; National Academies of Sciences, Engineering, and Medicine 2019; Niconchuk and Hyman 2020). The National Academies released a report in 2019, which synthesized the large body of literature on clinician burnout and proposed a multi-layered system-based model of the origins of burnout (National Academies of Sciences, Engineering, and Medicine 2019). They noted that interventions are needed at all levels of the healthcare system.

Mindfulness has emerged as a promising individual-level intervention that, in combination with system-level interventions, can improve professional well-being in clinicians (Shoker et al. 2024). Mindfulness involves paying purposeful attention to present-moment experiences with an attitude of acceptance and non-judgment (Kabat-Zinn 2003). This capacity is most often cultivated through mindfulness meditation (MM). Studies examining MM interventions among clinicians have demonstrated reductions in stress, anxiety, burnout, and emotional exhaustion (S. B. Goldberg et al. 2022; Irving, Dobkin, and Park 2009; Lamothe et al. 2016; Shoker et al. 2024). Our previous research found that self-reported mindfulness among GCs was negatively associated with burnout and compassion fatigue, and positively associated with empathy and work engagement (Silver et al. 2018). Thus, training mindfulness meditation in clinicians at risk for burnout may be an important strategy for retention and sustainable quality care in GCs.

Despite these promising findings, a significant limitation of mindfulness meditation research is the reliance on passive control groups, which fail to control for non-specific factors such as expectancy effects or structured self-care time (Davidson and Kaszniak 2015; S. B. Goldberg et al. 2017; S. Goldberg et al. 2019; Bishop 2002). Meditation trials with passive controls can overestimate effect sizes by 30-40% compared to those with active controls (Goyal et al. 2014; S. B. Goldberg et al. 2022). Inert active controls that mimic non-specific aspects of MM without incorporating mindfulness techniques are essential for accurately evaluating mindfulness’s unique contribution to outcomes. However, these have been challenging to design and no gold standard active control for MM exists (Davidson and Kaszniak 2015; Kinser and Robins 2013).

We sought to test the hypothesis that MM could improve professional well-being among GCs providing direct patient care in the United States. We designed a study comparing MM to both an active control meditation (ACM) and a no-meditation control (NMC) group. The ACM meditation was novel, informed by prior studies (Lindsay et al. 2019; Ruscio et al. 2016; Manocha et al. 2011) and designed to match the MM intervention in structure, duration, and frequency, while excluding explicit mindfulness techniques. Our primary aim was to test whether mindfulness meditation would be more effective than a well-matched active control meditation at reducing burnout among clinical GCs. The NMC group was included to allow comparison to a no-meditation passive control if the novel ACM arm did not perform well as an active control.

## METHODS

### Study Design

We conducted a double-blinded, parallel three-arm decentralized randomized controlled trial (RCT) with randomization 1:1:1 to three study arms: mindfulness meditation (MM), active control meditation (ACM), and no-meditation control (NMC). Participants were blinded to the study hypotheses. Individuals performing data analysis were blinded as to which arm participants were randomized. No protocol deviations or changes occurred. The primary outcome was burnout.

Secondary outcomes included stress, professional fulfillment, professional self-care, resilience, reactive distress, and a desire to spend less time caring directly for patients.

### Study Arms

#### Mindfulness meditation (MM) arm

The intervention was 8 weeks of 10 minutes a day of MM guided by Headspace, a web and smartphone app that provides MM exercises, guided by audio instructions from a teacher with extensive training in meditation practice and meditation instruction. Participants were asked to do specific courses in Headspace that included instruction on MM techniques like awareness of breath and sensations, noting of thinking or distraction, and gently returning attention to the breath. They were instructed to first do the Headspace Basics courses (30 sessions that facilitate learning and practice of the fundamentals of meditation and mindfulness) then, once finished Basics, to select from meditations that apply these fundamentals to managing anxiety, patience, restless, and navigating change (Radin et al. 2025). The intervention duration was chosen because most of the evidence for MM is from trials of 8-week interventions and sufficient evidence is not yet available for shorter intervention periods. Prior studies have shown that MM guided by Headspace can improve psychological symptoms in general (Zawadzki et al. 2025; Bostock et al. 2019; Flett et al. 2020), and professional well-being in healthcare workers specifically (Radin et al. 2025; Taylor et al. 2022).

#### Active control meditation (ACM) arm

Given that contemplative science currently lacks a gold standard control for MM, we developed a novel active control that was untested prior to this study. It was intended to be inert, meaning it did not include the active ingredient of mindfulness training. We created meditation exercises designed to mirror the structure (web-based, audio-guided), duration, and dose (10 minutes per day over 8 weeks) of MM, without mindfulness techniques. We drew from active control designs in prior studies on MM, where the control included elements of mind-wandering, introspection, and/or intentional thinking (Lindsay et al. 2019; Manocha et al. 2011; Ruscio et al. 2016). Audio instructions guided participants to close their eyes and get lost in their thoughts or reflect on their day, with no instruction regarding mindfulness (i.e., attention to breath and body sensations, acceptance of thoughts and feelings that arise, returning attention to the breath after noticing mind wandering to distraction). Mind wandering and getting lost in thoughts was chosen since it is distinct from mindfulness techniques which often encourage awareness of thought and non-judgmental observation of them, without getting lost in them or engaged with them, as well as returning attention to body sensations or other present-moment anchors once aware that attention has drifted to thoughts. Since this active control was novel, we included manipulation checks (change in mindfulness) and assessments of its credibility and expectancy (described below) to help assess its performance as an active control.

#### No-meditation control (NMC) arm

Participants were asked not to engage in any meditation exercises. NMC was included for secondary comparisons to MM in case ACM did not perform well as an active control.

### Sample Size Determination

The target sample size was determined based on a power analysis conducted for the primary outcome, burnout, measured via the Professional Fulfillment Inventory (PFI), comparing MM to ACM. To detect a 0.5 standard deviation difference in burnout scores, at a significance level of p < 0.05 and with 80% power, 213 participants are needed. We increased this to 387 to account for an expected lost-to-follow-up rate of 25%, lack of published data on PFI scores in this population, and an estimate that ∼15% of our sample would have a regular meditation practice and thus may not respond to the intervention (Silver et al. 2018).

### Participants

Recruitment occurred from September 2019 to July 2020 via NSGC e-blasts, NSGC forum posts, Twitter, and emails to participants in a prior study on this topic by our group (Silver et al., 2018). Recruitment materials noted that the study was on poor professional well-being, including burnout, but did not mention mindfulness as the intervention. Eligibility criteria included self-described English proficiency and being a GC in the United States who counsels patients. The study was limited to GCs who counsel patients because of secondary outcomes related to direct patient care, not reported here.

### Study procedures

Study procedures are outlined in Figure 1 and the CONSORT flow diagram is in Figure 2. This was a decentralized trial with participants completing all study procedures online.

**Figure 1.**
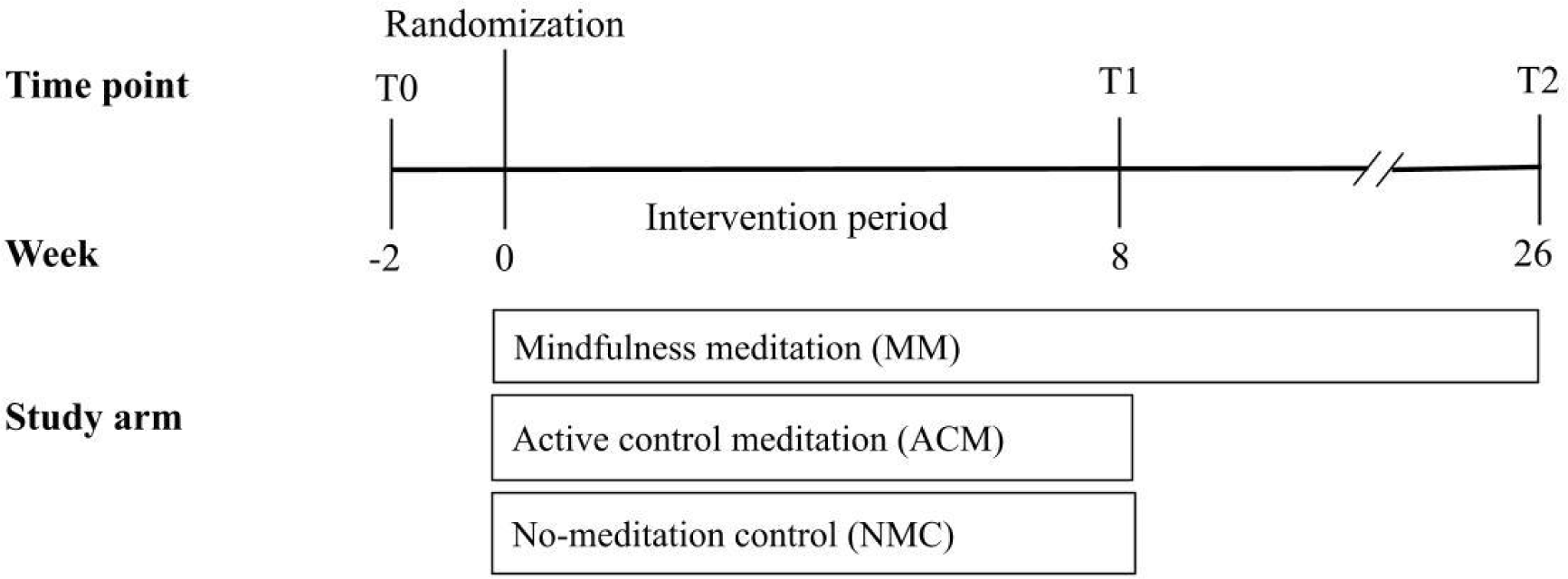
Study Procedures. Timeline and flow of study procedures for the three-arm randomized controlled trial. Participants completed the T0 (baseline) survey then were randomized 1:1:1 (mindfulness meditation, active control, no-meditation control). After the 8-week intervention period, participants were sent the T1 (post-intervention) survey. Participants in the mindfulness meditation arm completed an additional survey (T2) 26 weeks after randomization to assess maintenance of intervention effects.

**Figure 2.**
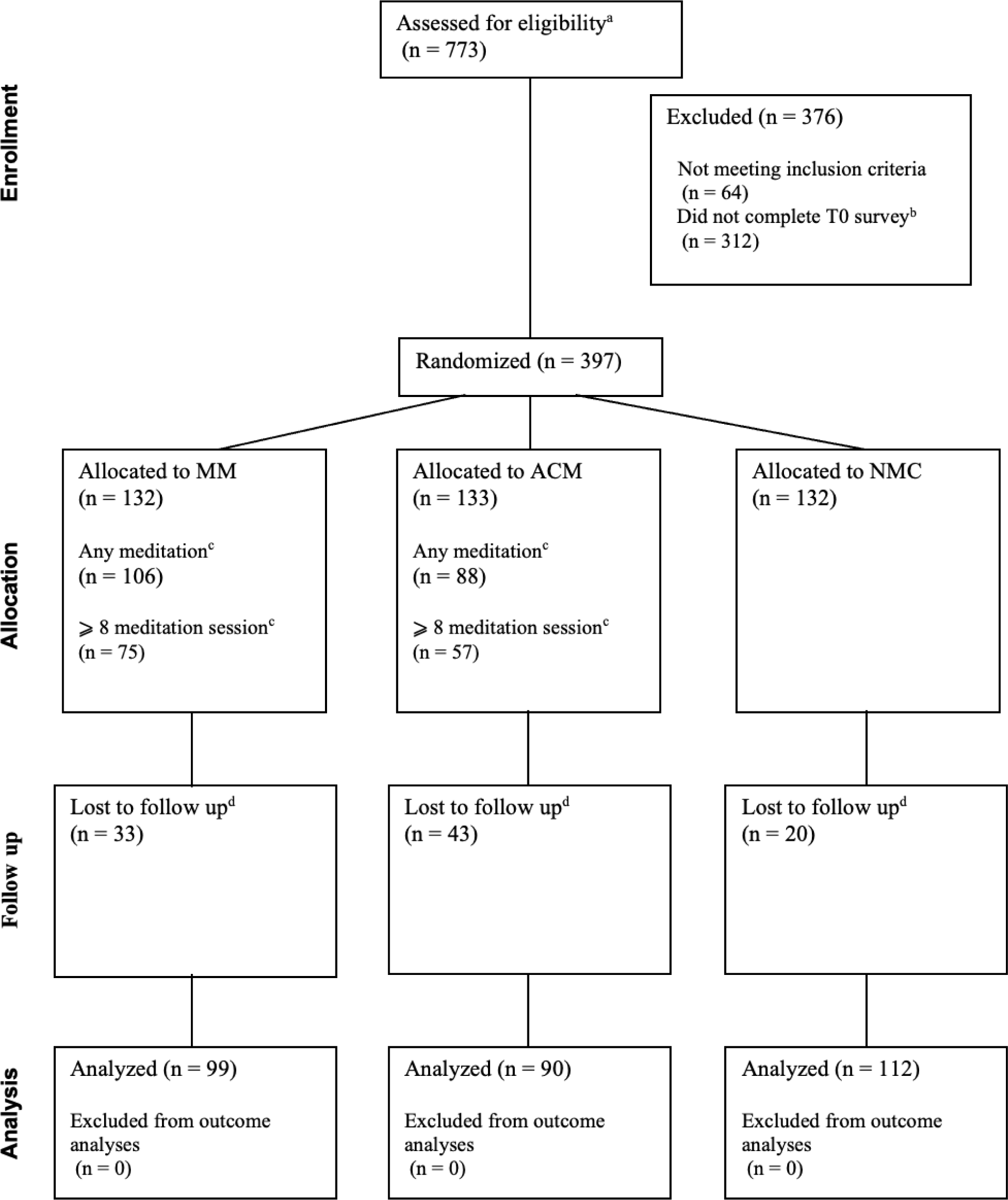
CONSORT diagram depicting participants’ flow through the trial. Flow of participants through screening, randomization, intervention, and follow-up phases of the three-arm randomized controlled trial. MM = mindfulness meditation, ACM = active control meditation, NMC = no-meditation control. ^a^Number of online eligibility surveys completed. The number of unique individuals who completed the eligibility survey may be less than 773 due to some individuals filling it out more than once. ^b^In order to be randomized, eligible participants had to complete the T0 survey. ^c^Reasons participants did not meditate were not collected. Eight sessions was used as a cut off because in the most studied mindfulness intervention, Mindfulness-Based Stress Reduction, 8 meditation sessions would be equivalent to only doing meditation in the weekly group sessions. ^d^Participants were considered lost to follow-up if they did not complete the T1 measure of burnout. Reasons for not completing that measure were not collected.

#### Randomization

Participants were randomized after completing the baseline (T0) survey so that randomization did not bias their T0 responses. Blocked randomization was used in order to increase the likelihood of balanced study group sizes. The randomization sequence was generated by the study statistician and randomization was assigned using the randomization module in REDCap.

#### Data Collection

Participants completed a survey at baseline (T0) and again after the 8-week intervention period (T1). MM participants only also completed a follow-up survey (T2) 26 weeks after randomization (Figure 1). Surveys were administered via REDcap.

### Performance of ACM

Given the novel nature of the ACM arm and the known challenges in designing active controls for meditation, we collected data to help assess how well the ACM meditation performed as an active control. This included credibility and expectancy, change in mindfulness from T0 to T1, and adherence to meditation instructions.

### Instrumentation

Supplemental Table 1 lays out the scales used, their properties (length, reliability, internal consistency), and when each was administered. Brief descriptions of each are provided below. Internal consistency of all scales in the present sample was assessed using Cronbach’s α (Supplemental Table 2).

Mindfulness: The Five-Factor Mindfulness Questionnaire (FFMQ) is a trait measure of dispositional mindfulness, developed using input from other existing mindfulness measures (Baer et al., 2006). It is sensitive to change with interventions and has been used in many studies of MM.

Credibility and Expectancy: The Credibility and Expectancy Questionnaire (CEQ) evaluates how believable participants find the intervention and their expectation of benefit. It is widely used in behavioral intervention trials to assess non-specific effects such as placebo or expectancy (Devilly & Borkovec, 2000).

Adherence to Meditation: For both ACM and MM, data on initiation of meditation sessions was collected via the platform that delivered the meditation. At T1, MM and ACM participants’ adherence to meditation instructions was assessed. We used investigator-created Likert-based questions to assess how often participants practiced techniques specific to each arm (e.g., focus on breath for MM and mind-wandering for ACM) then calculated a score for adherence to each type of meditation (MM, ACM). An open-ended question asked participants to describe what they did during meditation.

Perceived harms and benefits: At T1, MM and ACM participants were asked if they experienced any harms or negative effects, or any benefits and, in an open ended question, what those were.

Prior experience with meditation and mindfulness: At T0, participants were asked if they’d undergone prior meditation training and how often they meditate. At T1, participants were asked if they had prior mindfulness training.

Investigator-created questions were used to measure demographics (T1).

#### Primary outcome

Burnout: We used the burnout subscale of the PFI, which consists of the work exhaustion and interpersonal disengagement subscales (Trockel et al., 2018). The PFI is a relatively new measure of clinician well-being that was designed for sensitivity to change due to interventions. It is briefer than other measures of professional well-being yet maintains convergent validity with them (Trockel et al. 2018).

#### Secondary outcomes

Professional fulfillment: Professional fulfillment was measured using the professional fulfillment subscale of the PFI (Trockel et al., 2018).

Stress: The Perceived Stress Scale (PSS) is a measure of stress that has been extensively used in intervention studies, especially those targeting stress reduction (Cohen et al., 1983).

Professional self-care: The Professional Self-Care Scale (PSCS) measures the extent to which professionals engage in activities that may support their well-being. Subscales include professional support, professional development, life balance, cognitive awareness, and daily balance (Dorociak et al. 2017).

Resilience: The Connor-Davidson Resilience Scale (CD-RISC) evaluates personal qualities and resources that help individuals adapt to adversity. It is widely used to measure resilience in clinicians and other populations (Connor & Davidson, 2003).

Reactive distress: Reactive distress refers to discomfort and distress experienced in response to another person’s negative experience. It was measured using the personal distress subscale of the Interpersonal Reactivity Index (IRI), which assesses an individual’s tendency to experience distress in response to observing others’ difficult experiences (Davis, 1983).

Desire to reduce clinical time: This was assessed using an investigator-created question with a Likert scale that asked participants if they would like to reduce the amount of time they spend providing clinical care, including all aspects of clinical care.

### Data Analysis

We used a pre-specified intention-to-treat analysis for the primary outcome and all secondary outcomes. Outcomes were analyzed using a linear regression with the T1 score for each outcome as the dependent variable, study arm as a predictor, and the T0 score for the outcome as a covariate. T0 mindfulness (FFMQ) was also included in the model to control for dispositional or previously developed mindfulness, which has been found to moderate the effects of mindfulness interventions (Sieder et al. 2024). For each outcome, three pairwise comparisons were performed: MM vs ACM (primary comparison), MM vs NMC, and ACM vs NMC. While intervention effects were assessed via linear regression, we also report the following unadjusted analyses for descriptive purposes and in accordance with CONSORT: Differences between groups in T1 means of outcome variables, means of each outcome for each group at each timepoint, and Cohen’s d. As predefined in the protocol, we used multiple imputation to handle the missing T1 outcome data because >5% of values were missing. Multiple imputation was performed using the mice package in R using predictive mean matching with imputation of five datasets. The imputation model included randomization arm, baseline burnout, baseline mindfulness, and relevant demographic variables to address observed associations with being lost to follow-up. Convergence was verified by examining trace plots (which showed stable chain mixing and no trends across iterations), and the distributions of imputed values were similar to those of observed data.

We performed inductive coding and quantitative content analysis on open-ended responses. We used an alpha level of 0.05 for all statistical tests. Secondary outcomes were not included in sample size calculations and were analyzed without adjustment for multiple testing; results should be interpreted as exploratory and hypothesis-generating.

Reporting of this trial follows CONSORT 2025 guidelines (Supplemental Table 10; (Hopewell et al. 2025). No patients or members of the general public were involved in the trial design, conduct, or reporting. However, GCs led and contributed to the design, conduct, analysis, and reporting of this trial (CC, MAC, JA, JS). Study materials were piloted with multiple GCs prior to implementation. The study protocol is available upon request. The statistical analysis plan is described in the Data Analysis section above.

## RESULTS

### Participants

A total of 397 participants were randomized (132 MM, 133 ACM, 132 NMC). The mean age was 33.1 (SD=7.5), and nearly all participants were female, White, and non-Hispanic/Latine (Table 1). Most participants (64.5%) had some prior exposure to meditation; however, only a minority of participants (8.3%) meditated on a weekly basis or had some prior mindfulness training (11.7%; Supplemental Table 3).

**Table 1.** Participant characteristics.

|  | **Total** | **MM** | **ACM** | **NMC** |
| --- | --- | --- | --- | --- |
| Age<sup>a</sup> | 31.0 (10.0) | 31.0 (8.5) | 31.0 (10.0) | 31.0 (10.0) |
| Gender<sup>b</sup> |  |  |  |  |
| Female | 388/396 (98.0%) | 127/131 (96.9%) | 131/133 (98.5%) | 130/132 (98.5%) |
| Male | 8/396 (2.0%) | 4/131 (3.1%) | 2/133 (1.5%) | 2/132 (1.5%) |
| Race<sup>c</sup> |  |  |  |  |
| White | 374/397 (94.2%) | 125/132 (94.7%) | 131/133 (98.5%) | 118/132 (89.4%) |
| Asian | 24/397 (6.0%) | 7/132 (5.3%) | 2/133 (1.5%) | 15/132 (11.4%) |
| Black or African American | 2/397 (0.5%) | 1/132 (0.8%) | 1/133 (0.8%) | 0/132 (0%) |
| Other | 4/397 (1.0%) | 4/132 (3.0%) | 0/133 (0%) | 0/132 (0%) |
| Hispanic or Latine | 7/396 (1.8%) | 2/132 (1.5%) | 2/132 (1.5%) | 3/132 (2.3%) |
| Year of Graduation<sup>d</sup> | 2015 (8) | 2014 (9) | 2015 (8.25) | 2014 (8) |

<sup>a</sup> Median (IQR)<sup>b</sup> Transgender and non-binary options were provided but not selected; Male and Female were the options used in the surveys when they were administered. We have retained them here to ensure we are reporting the terms participants selected, however, they are not the current standard of the journal for gender, which is Man and Woman<sup>c</sup> Choose all that apply<sup>d</sup> Median (inter-quartile range)

Figure 2 presents the CONSORT flow diagram of participant progression through the trial. A quarter of the participants (24.2%) did not complete T1 outcome measures. More participants were lost to follow-up in the ACM (32%) and MM (25%) groups than the NMC group (15%) (p = 0.04). Participants lost to follow-up had higher baseline burnout scores (p = 0.02) and lower baseline mindfulness (subscales: acting with awareness, p = 0.005, non-judging, p = 0.002; non-reactivity, p = 0.03). More participants were lost to follow-up among those who did not engage in any meditation sessions in both the MM (69.2% vs. 13.2%) and ACM (77.8% vs. 9.1%) arms compared to those who meditated at least once (p<0.001).

Of note, the study started in 2019 and continued through 2020. For 39% of participants, the T1 survey and at least a portion of the intervention period occurred during the early months of the coronavirus pandemic. Neither T0 burnout or T1 burnout were associated with completing the relevant survey during the pandemic.

### Adherence to and experience with meditation

#### Credibility and Expectancy

The Credibility Expectancy Questionnaire (CEQ) was completed by 78.9% (104/132) of MM participants and 63.9% (85/133) of ACM participants. ACM participants rated their form of meditation as less credible and held lower expectations for benefit than MM participants did for their form of meditation (credibility: p < 0.0001; expectancy: p < 0.0001).

#### Adherence

The median number of meditation sessions was 13 (IQR 30.8) for MM and 5 (IQR 16.0) for ACM (p= 0.0001). More MM participants initiated at least one meditation session compared to ACM participants (Supplemental Table 4). Participants in both arms reported a high level of adherence to their assigned meditation instructions (Supplemental Table 5, Supplemental Figure 1). Number of meditation sessions initiated was not correlated with either T1 burnout (MM: p=0.25; ACM: p=0.45) or T1 mindfulness (MM: p=0.06; ACM: p=0.19). At T1, 29.0% (29/100) of MM participants and 22.5% of (20/89) ACM participants reported they had done meditation during the intervention period other than what was prescribed to them in the trial. However, they did very few non-study meditation sessions (MM: median 1.0 (IQR 1.0); ACM: median 1.0 (IQR 0.25)).

#### Perceived Harms and Benefits

Harms or negative effects associated with meditation were similarly reported across groups (MM 12.0% vs. ACM 15.7%; p = 0.6). Among self-reported harms and negative effects, more participants reported difficulty with the assigned meditation type in the ACM group (20.0%) than in the MM group (1.0%; p = 0.001; Supplemental Table 6). This included anxiety, agitation, and the focus on thoughts being unhelpful. More MM participants (91.0%) perceived benefit from meditation during the study compared to ACM participants (69.3%; p < 0.001). More MM participants intended to continue to meditate after the trial (MM 88.0%, ACM 75.3%; p = 0.04).

#### Change in trait mindfulness

Mindfulness increased from T0 to T1 by 10.7% in MM participants and 12.0% in ACM participants (Supplemental Table 7). Of note, change in mindfulness in ACM participants was not associated with self-reported use of mindfulness techniques during meditation (p = 0.5).

### Primary outcome - Burnout

In the pre-specified primary analysis using linear regression to evaluate the intervention effect, controlling for baseline burnout and mindfulness, no intervention effect was observed for MM, compared to ACM. However, in pre-specified secondary analyses, compared to NMC, both MM and ACM resulted in decreases in burnout (Table 2, Figure 3). Unadjusted mean differences between groups at T1 were 0.28 (95% CI −1.5 to 2.1) for MM vs ACM, −4.0 (95% CI −5.9 to −2.2) for MM vs NMC, and −3.8 (95% CI −5.7 to −1.8) for ACM vs NMC. Effect sizes for change in burnout from T0 to T1 were large for MM (Cohen’s d = −0.84, 95% CI −1.1 to −0.6) and medium for ACM (d = −0.70, 95% CI −1.0 to −0.4). Similar findings were seen for both components of burnout (Figure 3, Supplemental Table 8). Mean T2 burnout scores for MM were lower than T0 (p<0.001) but higher than T1 (p=0.03), suggesting partial maintenance of MM benefit (Table 3).

**Figure 3.**
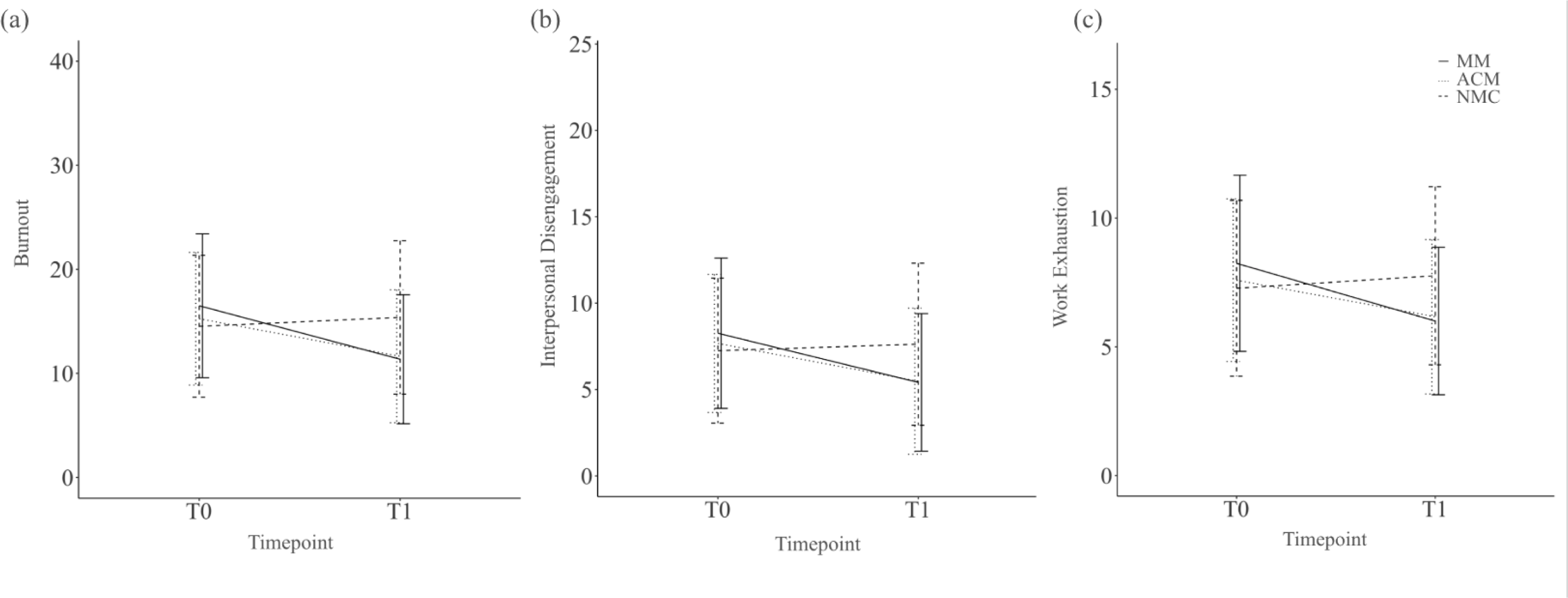
Burnout and its components by study arm and timepoint. Mean burnout scores and component subscales for each study arm at T0 (baseline) and T1 (after intervention). Error bars represent ± standard deviation from the mean. Panel (a) shows burnout subscal scores from the Professional Fulfillment Index. Panel (b) shows the interpersonal disengagement component of burnout. Panel (c) shows the work exhaustion component of burnout MM = mindfulness meditation, ACM = active control meditation, NMC = no-meditation control. The mindfulness meditation (MM) and active control meditation (ACM) groups both showed significant reductions in burnout compared to the no-meditation control (NMC) group, with no significant difference between MM and ACM groups. Sample sizes: MM n=100, ACM n=90, NMC n=111 at T1.

**Table 2.** Linear Regression Results for Intention-to-Treat Outcome Analysis for Burnout.

| **Comparison** | **Term** | **Coefficient ($\beta$)** | **Standard Error** | **p-value** |
| --- | --- | --- | --- | --- |
| MM vs ACM | Study Arm | -0.53 | 0.83 | 0.525 |
|  | T0 Burnout | 0.28 | 0.06 | <0.0001 |
|  | T0 Mindfulness | 0.02 | 0.08 | 0.812 |
| ACM vs NMC | Study Arm | -3.36 | 0.78 | <0.0001 |
|  | T0 Burnout | 0.49 | 0.06 | <0.0001 |
|  | T0 Mindfulness | -0.05 | 0.09 | 0.548 |
| MM vs NMC | Study Arm | -4.05 | 0.77 | <0.0001 |
|  | T0 Burnout | 0.44 | 0.06 | <0.0001 |
|  | T0 Mindfulness | -0.05 | 0.09 | 0.593 |

1. Abbreviations:
- MM: Mindfulness Meditation - ACM: Active Control Meditation - NMC: No-Meditation Control
2. Sample Size: N = 397

**Table 3.** Burnout at Each Timepoint, by Study Arm.

| **Timepoint** | **Arm** | **Mean (95% CI)** | **Burnt Out** |
| --- | --- | --- | --- |
| T0 | MM | 16.5 (15.3–17.7) | 75/131 (57.3%) |
|  | ACM | 15.2 (14.2–16.3) | 60/133 (45.5%) |
|  | NMC | 14.5 (13.3–15.7) | 80/132 (60.2%) |
| T1 | MM | 11.4 (10.1–12.6) | 35/100 (35.0%) |
|  | ACM | 11.6 (10.3–13.0) | 35/90 (38.8%) |
|  | NMC | 15.4 (14.0–16.8) | 66/111 (59.4%) |
| T2 | MM | 13.0 (11.4–14.5) | 37/87 (38.1%) |

1. Burnout Cutoff: A score $\geq 14$ indicates the likelihood of experiencing burnout, as defined by scale authors. 2. Abbreviations: - MM: Mindfulness Meditation - ACM: Active Control Meditation - NMC: No-Meditation Control

### Secondary outcomes

For perceived stress, compared to NMC, both MM and ACM resulted in greater reductions, with, no difference between MM and ACM (Supplemental Table 9, Supplemental Figure 2). Unadjusted mean differences in stress scores between groups at T1 were 1.2 (95% CI −0.5 to −3.0, p=0.16) for MM vs ACM, −3.7 (95% CI −5.3 to −2.0 p<0.001) for MM vs NMC, and −4.9 (95% CI −6.6 to −3.2, p<0.01) for ACM vs NMC. Effect sizes for change in stress from T0 to T1 were large for both MM (Cohen’s d=-0.88, 95% CI −1.16 to −0.60 and ACM (d=-0.90, 95% CI −1.19 to −0.60). Mean T2 stress scores for MM were comparable to T0 (p=0.4) and higher than T1 (p<0.001), suggesting a lack of sustained benefit. Of note, all participants completed the T2 survey during the first year of the coronavirus pandemic.

For the professional self-care scale, compared to NMC, MM resulted in significantly greater self-care. However, there was no difference for MM vs. ACM and for ACM vs NMC (Supplemental Table 9). The MM intervention effect was driven by increases in the cognitive awareness subscale. No other subscales showed intervention effects. T2 cognitive awareness subscale scores for MM participants were higher than T0 (p = 0.0001), and comparable to T1 (p = 0.1), suggesting that benefit was sustained. The effect size for change in cognitive awareness from T0 to T1 for MM was medium (Cohen’s d = 0.70, 95% CI 0.43 to 0.98).

No differences were observed between study arms for professional fulfillment, resilience, desire to reduce time spent seeing patients, and reactive distress (Supplemental Table 9).

## DISCUSSION

The primary aim of this study was to examine the effects of 8-weeks of mindfulness meditation training (MM) on self-reported burnout, as compared to an active control meditation (ACM) that did not include mindfulness training. Both MM and ACM yielded similar reductions in burnout, which was not what we expected. The non-superiority of MM vs ACM may be explained by factors related to the performance of the active control condition. ACM was designed to control for non-specific aspects of meditation—such as dedicated self-care time, structured practice, and expectation of benefit—while excluding explicit mindfulness training (Davidson and Kaszniak 2015; Kinser and Robins 2013). However, mindfulness increased similarly in both MM and ACM groups. Furthermore, lower credibility and expectancy scores suggest that ACM failed to control for the expectation of benefit, a critical component of a well-matched active control (Bishop, 2002).

Given these findings, the ACM failed to serve as an appropriate active control training, preventing a valid test of our primary hypothesis regarding the specific effects of mindfulness meditation versus non-specific meditation effects. In pre-planned secondary analyses, compared to a passive no-meditation control (NMC), MM yielded significantly increased mindfulness, decreased burnout, and decreased perceived stress.

The poor performance of ACM as an active control is consistent with broader challenges in meditation research regarding the design of appropriate control conditions and the lack of a gold standard active control for mindfulness meditation (Van Dam et al. 2018; Davidson and Kaszniak 2015; S. Goldberg et al. 2019). The unexpected increase in mindfulness among ACM participants reveals ACM was not in fact inert. However, Supplemental Figure 1 reveals that despite both arms showing increased mindfulness scores, participants were indeed engaging in fundamentally different practices. It may be that ACM inadvertently included mindfulness components (S. B. Goldberg et al. 2016; Tran et al. 2022). A 2022 meta-analysis of 146 RCTs found that self-reported mindfulness increased in (non-mindfulness) active controls and that change in mindfulness accounted for change in mental health outcomes (Tran et al. 2022). One possibility is that ACM instructions inadvertently promoted mindfulness through the cultivation of meta-awareness—the awareness of one’s current mental state, including where attention is focused, a key component of mindfulness (Jankowski and Holas 2014; Dunne, Thompson, and Schooler 2019). Although designed to encourage mind-wandering and reflection without mindfulness techniques, the ACM’s periodic audio prompts to “get lost in your thoughts” may have paradoxically increased participants’ awareness of their mental states. Additionally, the intentional nature of mind-wandering in ACM may differ from spontaneous mind-wandering. Research by Seli and colleagues found that intentional mind-wandering is positively associated with mindfulness as measured by the FFMQ, while unintentional mind-wandering shows negative associations (Seli, Carriere, and Smilek 2015).

Given the increase in mindfulness and decrease in burnout observed in the ACM arm, it is worth considering whether ACM may be an effective intervention for addressing GC burnout. Notably, multiple data points suggest that ACM is less acceptable than MM for at least a subset of GCs. In addition to having lower credibility and expectancy than MM, ACM had more participants that were lost to follow-up, more participants that did not meditate during the trial, fewer meditation sessions per participant, fewer participants who perceived meditation benefited them, and fewer participants who intended to continue to meditate after the trial. Only 1.0% of MM participants reported difficulty with their assigned meditation type, compared to 20.0% of ACM participants (p = 0.001). ACM participants described experiencing anxiety and agitation, and finding the focus on thoughts unhelpful—experiences that are both harmful and that could limit long-term adherence and effectiveness. These data suggest that the form of meditation used in the ACM may not be a viable option for many GCs.

Although our primary hypothesis could not be effectively tested, pre-planned secondary analyses provide evidence for MM’s positive impact on professional well-being in clinical GCs. Given the known challenges in designing active controls for MM, we included a passive control arm (NMC) to serve as an alternate control in secondary analyses in case ACM did not perform effectively as an inert active control. Compared to NMC, MM reduced both burnout and stress with large effect sizes. At baseline, 57.3% of MM participants were burnt out, dropping to 35.0% at T1. In contrast, the proportion of GCs experiencing burnout remained stable in the NMC arm. These findings need to be interpreted with caution, given that they arise from secondary analyses, and because it has been well demonstrated that comparisons to a passive control can both overestimate effect sizes and identify effectiveness erroneously (Davidson and Kaszniak 2015; Kinser and Robins 2013; S. B. Goldberg et al. 2022). However, recent active-controlled trials support the validity of the benefits of MM that we observed. Several randomized studies have demonstrated that mindfulness interventions outperform active comparators for clinician burnout and stress, including dedicated break time (Ireland et al. 2017), stress education (Cascales-Pérez et al. 2021), and protected time for self-directed activities (West et al. 2014). A 2022 systematic review found mindfulness superior to active controls for stress in healthcare providers (S. B. Goldberg et al. 2022). This convergent evidence from active-controlled studies suggests our observed intervention effect for MM versus NMC is less likely to be spurious and more likely to represent true benefit for clinical GCs.

We did not see a dose dependent relationship between number of meditation sessions and T1 burnout or mindfulness, which is consistent with prior studies (Strohmaier 2021). However, recent studies suggest a longer time frame may be needed to detect a relationship between meditation dose and benefit (Bowles, Davies, and Van Dam 2022; Bowles and Van Dam 2025). The fact that participants who did not complete the study had higher baseline burnout suggests that self-initiated app-based MM may not be accessible to GCs who are more severely burnt out. They may instead need more structured support such as MM facilitated in the work setting, peer-support, or systems-level interventions, which are discussed further below (M. Klatt, Steinberg, and Duchemin 2015; West et al. 2014; Panagioti et al. 2017).

Among secondary outcomes, MM reduced stress and increased scores on the cognitive awareness subscale of the Professional Self Care Scale, but it did not impact professional fulfillment, resilience, reactive distress, or desire to reduce clinical time. MM may not be a good fit for these outcomes or the trial may have been underpowered to detect an intervention effect for them.

GCs often have access to MM resources provided by their employers. This can include classes such as Mindfulness-Based Stress Reduction, apps such as Headspace and Calm, or brief recurring guided MM sessions in the workplace. Studies across a variety of work settings have found investment in mindfulness programs leads to increased productivity and reduced healthcare usage by employees, which can help make the business case for employers to provide such programs (Huberty et al. 2022; M. D. Klatt et al. 2016). A recent qualitative meta-synthesis examined factors that affect successful implementation of MM for healthcare providers (Knudsen et al. 2024). The authors provide recommendations including, but not limited to, leadership support, providing protected time during work and dedicated space, making participation optional, offering group-based trainings, providing training in brief exercises that can be done during clinical work, and providing in-person and online options. In terms of feasibility, it is notable that our results and those from other studies suggest that the benefits of MM can be realized with short (∼10 minutes) meditation sessions done a few times a week (Bostock et al. 2019; Fincham, Mavor, and Dritschel 2023; Champion, Economides, and Chandler 2018). In addition, self-directed app-based meditation, which can be more accessible to busy clinicians than live classes, has been shown to be effective for a variety of outcomes, including stress, burnout, and other aspects of professional well-being (Zawadzki et al. 2025; Bostock et al. 2019; Flett et al. 2020; Radin et al. 2025; Taylor et al. 2022; Champion, Economides, and Chandler 2018; Gál, Ștefan, and Cristea 2021).

Even when time and resources for MM are provided by an employer, MM is still ultimately an individual-level intervention. Focusing solely on individual-level interventions is insufficient to effectively address clinician burnout and is also unethical (De Simone, Vargas, and Servillo 2021; Pijpker et al. 2019; Spilg 2024; National Academies of Sciences, Engineering, and Medicine 2019). This is evident in our own data, with a third of MM participants still experiencing burnout at T1. As articulated in the National Academy of Medicine’s report on clinician burnout (National Academies of Sciences, Engineering, and Medicine 2019), burnout is fundamentally a systems-level problem requiring systems-level solutions. Meta-analyses and systematic reviews have found that work-system interventions have larger effects on clinician burnout than individual-level interventions (De Simone, Vargas, and Servillo 2021; Panagioti et al. 2017) and that combinations of the two are most effective (Pijpker et al. 2019). Our previous work identified multiple work-system factors contributing to GC burnout, including insufficient administrative support, lack of autonomy, and not feeling valued by non-GC colleagues (Caleshu et al. 2022). The broader literature on clinician burnout suggests additional work-system interventions including a culture where clinicians can speak up and have meaningful involvement in organizational decisions that affect them, improvements in the usability of technology, ensuring workload aligns with resources, peer-support programs that enhance sense of community and meaning at work, reduction in documentation burden, and increased automation of low-in-scope tasks (De Simone, Vargas, and Servillo 2021; Thomas Craig et al. 2021; Pijpker et al. 2019; National Academies of Sciences, Engineering, and Medicine 2019).

### Limitations

Several limitations warrant consideration when interpreting our findings. Since participants who were lost to follow-up had higher burnout and lower mindfulness at baseline, outcome data was unlikely to be missing completely at random, which is a key assumption of multiple imputation. However, we included these baseline variables in the imputation model, which can mitigate bias. Convergence was verified by examining trace plots (which showed stable chain mixing and no trends across iterations), and the distributions of imputed values were similar to those of observed data, suggesting multiple imputation was appropriate in this context. Our measurement of meditation adherence may overestimate actual practice, as the modalities available for collecting this data could only measure session initiation, not completion. Additionally, 39% of participants completed their post-intervention assessment during the early months of the COVID-19 pandemic. While we found no association between pandemic timing and burnout, the unprecedented stressors of this period may have influenced results in unmeasured and/or unmeasurable ways. While we aimed to blind both participants and analysts, this likely wasn’t fully effective. The daily balance subscale of the Professional Self Care Scale had low internal consistency in our sample. Our participants were nearly all White, cis-gender, and female. This homogeneity limits generalizability to GCs of other identities, who may have different experiences of both burnout and meditation. Future research must prioritize recruiting diverse samples to understand whether meditation’s benefits extend equally across all GCs.

### Future Directions

In addition to the need for studies on GCs of varying identities, research on MM for GC students and GCs not involved in direct patient care are also needed. Further work is needed to develop and validate active controls that successfully control for non-specific aspects of meditation without inducing mindfulness. This methodological work is essential for determining mindfulness meditation’s specific versus non-specific benefits (S. B. Goldberg et al. 2022). Studies on clinician mindfulness meditation have found improvement in care quality and patient outcomes; investigating this within the genetic counseling field would be valuable (Braun, Kinser, and Rybarczyk 2019). Lastly, studies are needed on a range of work system-level interventions to improve GC burnout and other aspects of professional well-being (National Academies of Sciences, Engineering, and Medicine 2019).

### Conclusion

Our primary hypothesis could not be effectively evaluated due to methodological challenges with the active control. Nonetheless, this randomized controlled trial provides evidence that meditation can significantly reduce burnout and stress among GCs. Pre-planned secondary analyses showed that MM improved burnout and stress in clinical GCs when compared to a passive control. These findings, when considered within the context of the mounting evidence for positive impact of clinician meditation on burnout, point to the need to consider MM as an individual-level intervention to mitigate burnout in the genetic counseling field. However, meditation must be positioned as one component of a comprehensive approach to GC professional well-being, complementing rather than replacing necessary systems-level reforms.

## AUTHOR CONTRIBUTIONS

Author contributions are described in alignment with CONSORT and CReDiT (Hopewell et al. 2025; National Information Standards Organization (U.S.) 2005). CC, MAC, JA, PG, JS and AT were responsible for conceptualization; CC was responsible for funding acquisition with input from MAC, JA, and AT; CC, MAC, JA, PG, and AT were responsible for methodology; CC was responsible for investigation, project administration, data curation, formal analysis, visualization, and writing – original draft; MAC, JA, and AT provided supervision; all authors contributed to writing – review & editing. All authors critically revised’ the manuscript for important intellectual content and approved the final version. CC is the guarantor of this work. The corresponding author attests that all listed authors meet authorship criteria and that no others meeting the criteria have been omitted.

## Data Availability

All data produced in the present study are available upon reasonable request to the authors

## ACKNOWLEDGEMENTS

ChatGPT and Claude were used throughout the manuscript. Specifically, drafting text based on human input, suggesting revisions to text drafted by humans, and drafting tables based on provided data. In all instances where generative AI tools were used, a human author reviewed and revised the tool’s output before incorporation into the manuscript. The Stanford REDCap platform was used in the trial and (http://redcap.stanford.edu) is developed and operated by the Stanford Medicine Research Technology team. The REDCap platform services at Stanford are subsidized by a) Stanford School of Medicine Research Office, and b) the National Center for Research Resources and the National Center for Advancing Translational Sciences, National Institutes of Health, through grant UL1 TR003142 (Harris et al. 2019). The study was funded by the Jane Engelberg Memorial Fellowship. Headspace provided in-kind six-month app subscriptions to all study participants. We thank Alex McMillan, Ondrej Blaha, and Kristopher Kapphahn for consultative statistical support, and Alyssa Schweickert and Isabela Dall Oglio Bucco for administrative support. This work was conducted to fulfill degree requirements (CC, PhD candidate at Leiden University Medical Center).

---

**Supplemental Figure 1:**
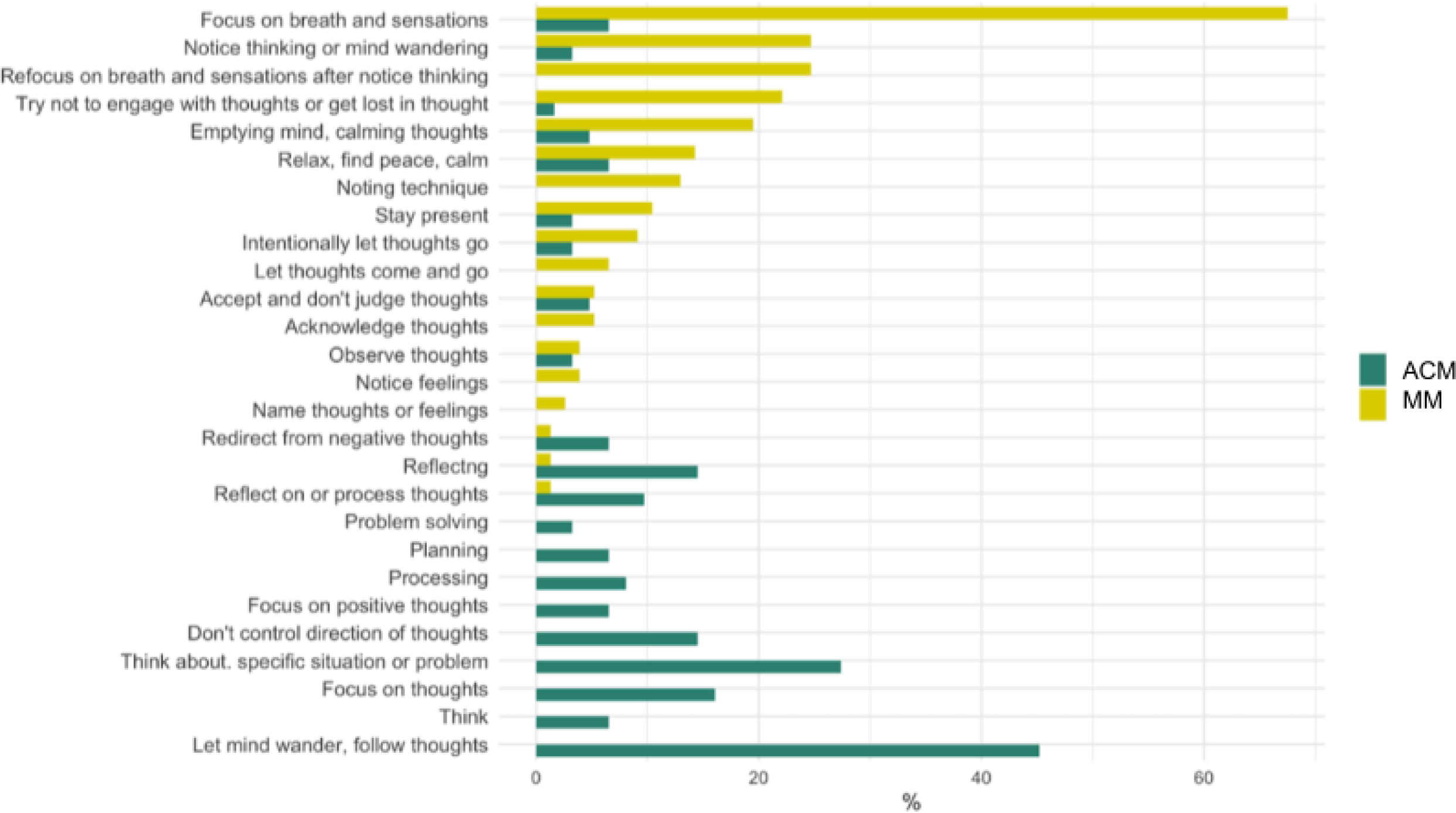
Participant descriptions of what they did while meditating. Frequency of meditation techniques reported by participants in open-ended responses describing their meditation practice during the final week of the intervention. Yellow bars represent mindfulness meditation (MM) participants (n=77); teal bars represent active control meditation (ACM) participants (n=62). Data analyzed using quantitative content analysis and inductive coding demonstrate that participants in each arm engaged in fundamentally different practices consistent with their assigned meditation type, suggesting participants understood and followed the meditation instructions they were randomized to.

**Supplemental Figure 2.**
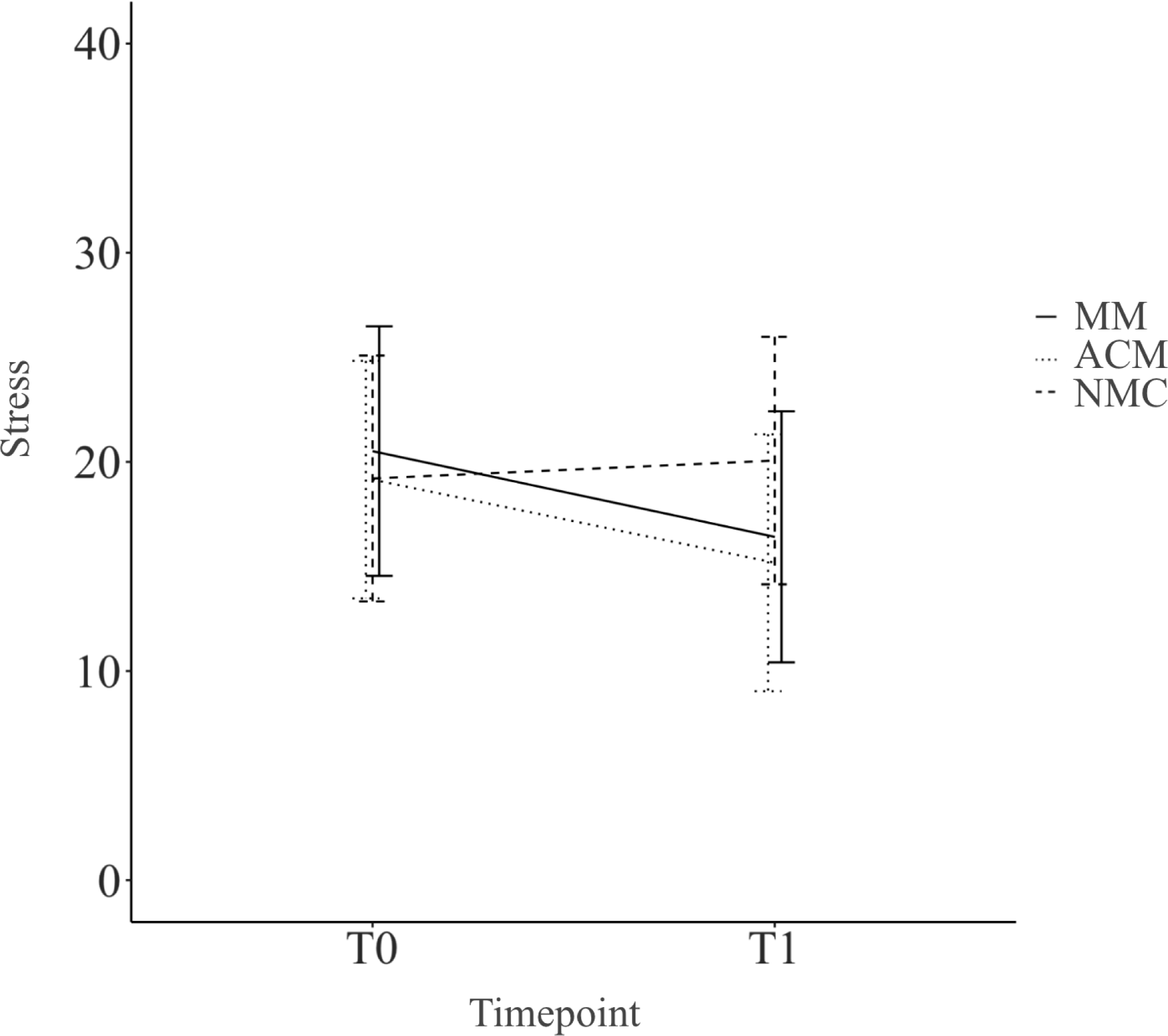
Stress by study arm and timepoint. Mean perceived stress scores for each study arm at baseline (T0) and post-intervention (T1), measured using the Perceived Stress Scale. Error bars represent ± one standard deviation from the mean. Both mindfulness meditation (MM) and active control meditation (ACM) groups showed significant stress reduction compared to the no-meditation control (NMC) group, with no significant difference between meditation groups. Sample sizes: MM n=100, ACM n=90, NMC n=111 at T1.

**Supplemental Table 1.**
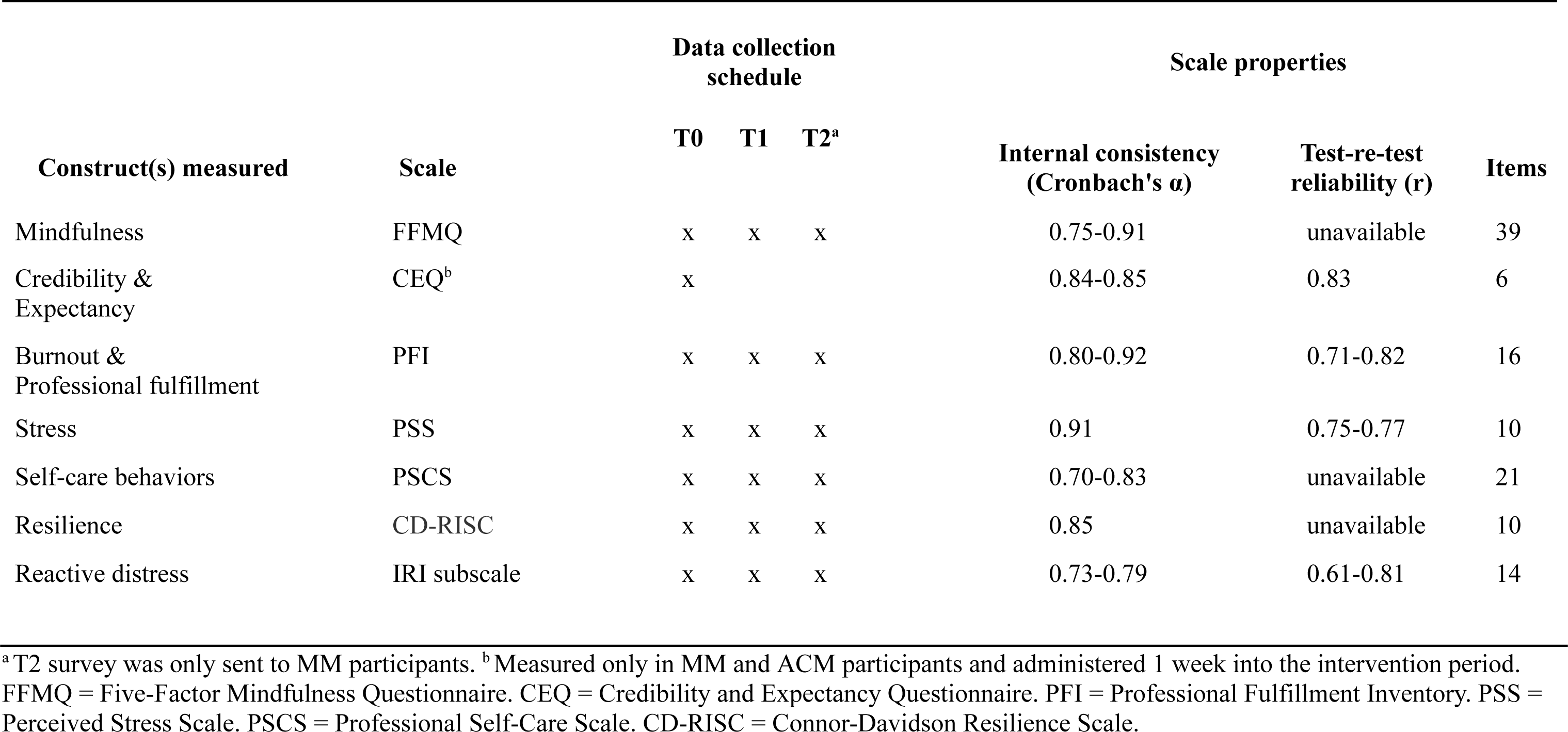
Properties and timing of scales used.

| Construct(s) measured | Scale | Data collection schedule |  |  | Scale properties |  |  |
| --- | --- | --- | --- | --- | --- | --- | --- |
| | | T0 | T1 | T2<sup>a</sup> | Internal consistency (Cronbach's $\alpha$) | Test-re-test reliability (r) | Items |
| Mindfulness | FFMQ | x | x | x | 0.75-0.91 | unavailable | 39 |
| Credibility & Expectancy | CEQ<sup>b</sup> | x |  |  | 0.84-0.85 | 0.83 | 6 |
| Burnout & Professional fulfillment | PFI | x | x | x | 0.80-0.92 | 0.71-0.82 | 16 |
| Stress | PSS | x | x | x | 0.91 | 0.75-0.77 | 10 |
| Self-care behaviors | PSCS | x | x | x | 0.70-0.83 | unavailable | 21 |
| Resilience | CD-RISC | x | x | x | 0.85 | unavailable | 10 |
| Reactive distress | IRI subscale | x | x | x | 0.73-0.79 | 0.61-0.81 | 14 |

<sup>a</sup> T2 survey was only sent to MM participants. <sup>b</sup> Measured only in MM and ACM participants and administered 1 week into the intervention period. FFMQ = Five-Factor Mindfulness Questionnaire. CEQ = Credibility and Expectancy Questionnaire. PFI = Professional Fulfillment Inventory. PSS = Perceived Stress Scale. PSCS = Professional Self-Care Scale. CD-RISC = Connor-Davidson Resilience Scale.

**Supplemental Table 2.** Cronbach’s Alpha (α) for Scales by Study Arm.

| Scale | Full Cohort (N) | ACM (N) | NMC (N) | MM (N) |
| --- | --- | --- | --- | --- |
| **FFMQ** |  |  |  |  |
| Observing | 0.79 (394) | 0.77 (132) | 0.77 (132) | 0.84 (130) |
| Describing | 0.90 (391) | 0.89 (131) | 0.90 (130) | 0.90 (130) |
| Acting with Awareness | 0.88 (390) | 0.87 (132) | 0.88 (130) | 0.90 (128) |
| Nonjudging of Inner Experience | 0.94 (391) | 0.94 (130) | 0.94 (132) | 0.95 (129) |
| Nonreactivity to Inner Experience | 0.83 (394) | 0.84 (130) | 0.81 (132) | 0.85 (132) |
| **CEQ** |  |  |  |  |
| Credibility | 0.86 (177) | 0.87 (84) | — | 0.80 (93) |
| Expectancy | 0.83 (176) | 0.83 (84) | — | 0.80 (92) |
| **PFI** |  |  |  |  |
| Work Exhaustion | 0.84 (396) | 0.80 (133) | 0.85 (132) | 0.87 (131) |
| Interpersonal Disengagement | 0.87 (393) | 0.85 (132) | 0.88 (132) | 0.88 (129) |
| Burnout (composite) | 0.89 (393) | 0.88 (132) | 0.90 (132) | 0.90 (129) |
| Professional Fulfillment | 0.88 (393) | 0.87 (131) | 0.87 (132) | 0.88 (130) |
| **PSS** |  |  |  |  |
|  | 0.87 (395) | 0.85 (131) | 0.87 (132) | 0.88 (132) |
| **PSCS** |  |  |  |  |
| Total Score | 0.87 (382) | 0.87 (129) | 0.86 (127) | 0.88 (126) |
| Professional Support | 0.81 (396) | 0.82 (132) | 0.79 (132) | 0.82 (132) |
| Professional Development | 0.68 (392) | 0.67 (132) | 0.71 (130) | 0.66 (130) |
| Life Balance | 0.84 (392) | 0.87 (133) | 0.84 (129) | 0.78 (130) |
| Cognitive Awareness | 0.69 (392) | 0.70 (131) | 0.69 (131) | 0.69 (130) |
| Daily Balance | 0.44 (395) | 0.51 (133) | 0.28 (130) | 0.53 (132) |
| **CD-RISC** |  |  |  |  |
|  | 0.83 (393) | 0.82 (131) | 0.79 (131) | 0.87 (131) |
| **IRI** |  |  |  |  |
| Personal Distress | 0.80 (392) | 0.81 (132) | 0.76 (129) | 0.83 (131) |

Cronbach's $\alpha$ values represent internal consistency for each scale or subscale within cohort groups at T0. ACM = Active Control Meditation; NMC = No-Meditation Control; MM = Mindfulness Meditation. FFMQ = Five-Factor Mindfulness Questionnaire. CEQ = Credibility and Expectancy Questionnaire. PFI = Professional Fulfillment Inventory. PSS = Perceived Stress Scale. PSCS = Professional Self-Care Scale. CD-RISC = Connor-Davidson Resilience Scale.

**Supplemental Table 3.**
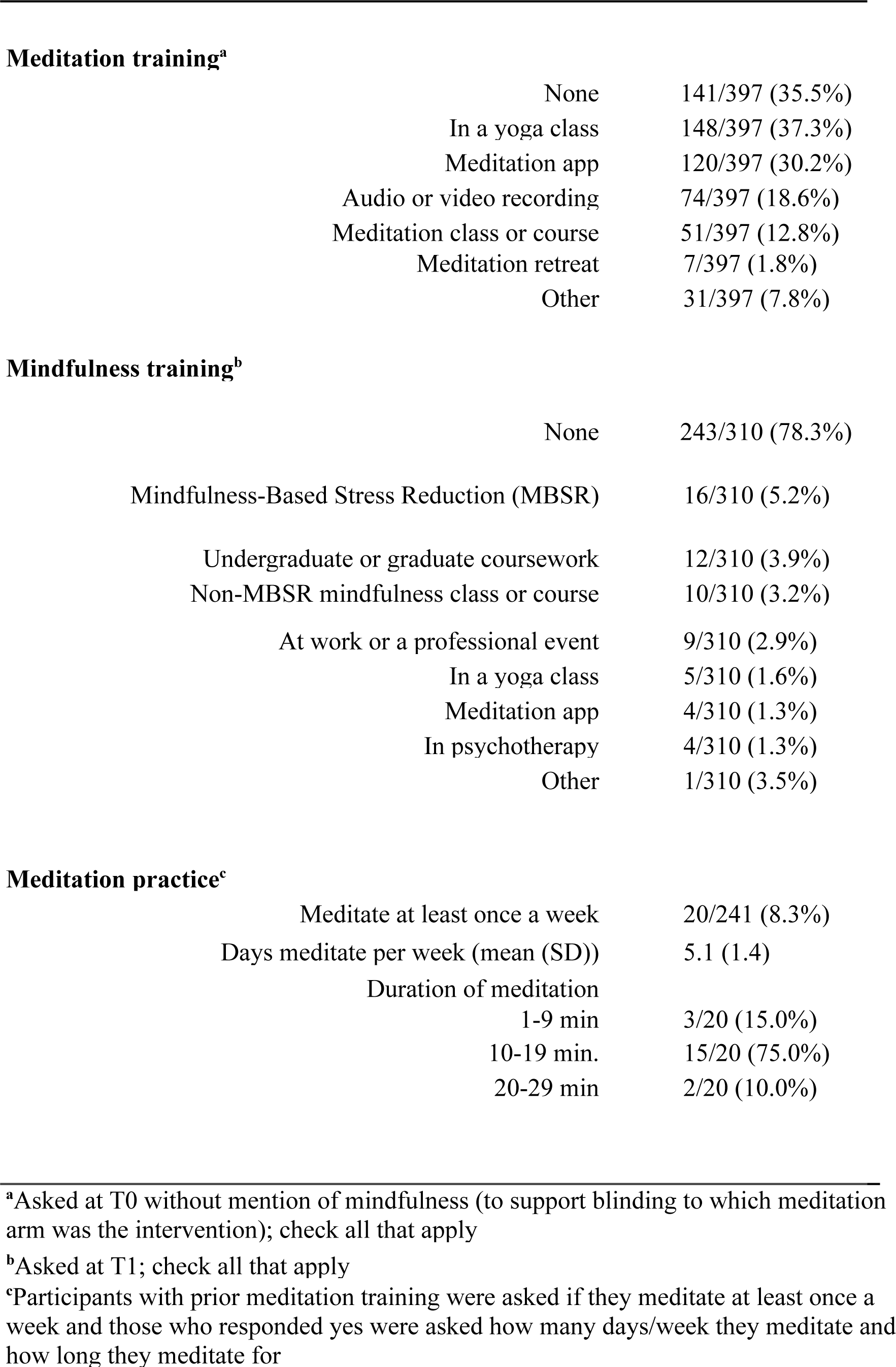
Prior experience with meditation and mindfulness.

**Meditation training<sup>a</sup>**
|  |  |
| --- | --- |
| None | 141/397 (35.5%) |
| In a yoga class | 148/397 (37.3%) |
| Meditation app | 120/397 (30.2%) |
| Audio or video recording | 74/397 (18.6%) |
| Meditation class or course | 51/397 (12.8%) |
| Meditation retreat | 7/397 (1.8%) |
| Other | 31/397 (7.8%) |
| **Mindfulness training<sup>b</sup>** |  |
| None | 243/310 (78.3%) |
| Mindfulness-Based Stress Reduction (MBSR) | 16/310 (5.2%) |
| Undergraduate or graduate coursework | 12/310 (3.9%) |
| Non-MBSR mindfulness class or course | 10/310 (3.2%) |
| At work or a professional event | 9/310 (2.9%) |
| In a yoga class | 5/310 (1.6%) |
| Meditation app | 4/310 (1.3%) |
| In psychotherapy | 4/310 (1.3%) |
| Other | 1/310 (3.5%) |
| **Meditation practice<sup>c</sup>** |  |
| Meditate at least once a week | 20/241 (8.3%) |
| Days meditate per week (mean (SD)) | 5.1 (1.4) |
| Duration of meditation |  |
| 1-9 min | 3/20 (15.0%) |
| 10-19 min. | 15/20 (75.0%) |
| 20-29 min | 2/20 (10.0%) |

<sup>a</sup>Asked at T0 without mention of mindfulness (to support blinding to which meditation arm was the intervention); check all that apply
<sup>b</sup>Asked at T1; check all that apply
<sup>c</sup>Participants with prior meditation training were asked if they meditate at least once a week and those who responded yes were asked how many days/week they meditate and how long they meditate for

**Supplemental Table 4.** Meditation Adherence.

| Adherence Metric | MM (n=132) | ACM (n=133) | p-value |
| --- | --- | --- | --- |
| Any meditation sessions, n (%) | 106 (80.3%) | 88 (66.2%) | 0.009 |
| Meditation dose (all participants)<sup>a</sup> | 13.0 (30.8) | 5.0 (16.0) | 0.0001 |
| Meditation dose (among those who meditated)<sup>a</sup> | 17.0 (30.8) | 12.0 (17.2) | 0.004 |

<sup>a</sup> Median number of sessions initiated (IQR)
1. Abbreviations:
- MM: Mindfulness Meditation - ACM: Active Control Meditation - NMC: No-Meditation Control - IQR: Inter-Quartile Range

**Supplemental Table 5.** Adherence to Meditation Instructions.

| Instruction Type | MM (n=100) | ACM (n=90) | p-value |
| --- | --- | --- | --- |
| Mindfulness techniques score<sup>a</sup> (mean (SD)) | 14.9 (4.2) | 9.0 (3.8) | <0.001 |
| Active control techniques score<sup>a</sup> (mean (SD)) | 10.1 (3.9) | 14.8 (4.1) | <0.001 |
| Self-reported mindfulness techniques<sup>b</sup> (n (%)) | 63/77 (81.8%) | 7/62 (11.3%) | <0.001 |
| Self-reported ACM techniques<sup>#</sup> (n (%)) | 2/77 (2.6%) | 43/62 (69.4%) | <0.001 |

<sup>a</sup> Scores range from 0-20, with higher scores indicating better adherence to the respective meditation type's specific instructions based on Likert-scale responses to 5 questions per meditation type
<sup>b</sup> Based on quantitative content analysis of open-ended responses describing meditation practice; participants were coded as using either MM or ACM techniques if they mentioned at least one technique specific to that meditation type
1. Abbreviations:
- MM: Mindfulness Meditation - ACM: Active Control Meditation - NMC: No-Meditation Control - SD: Standard Deviation

**Supplemental Table 6.**
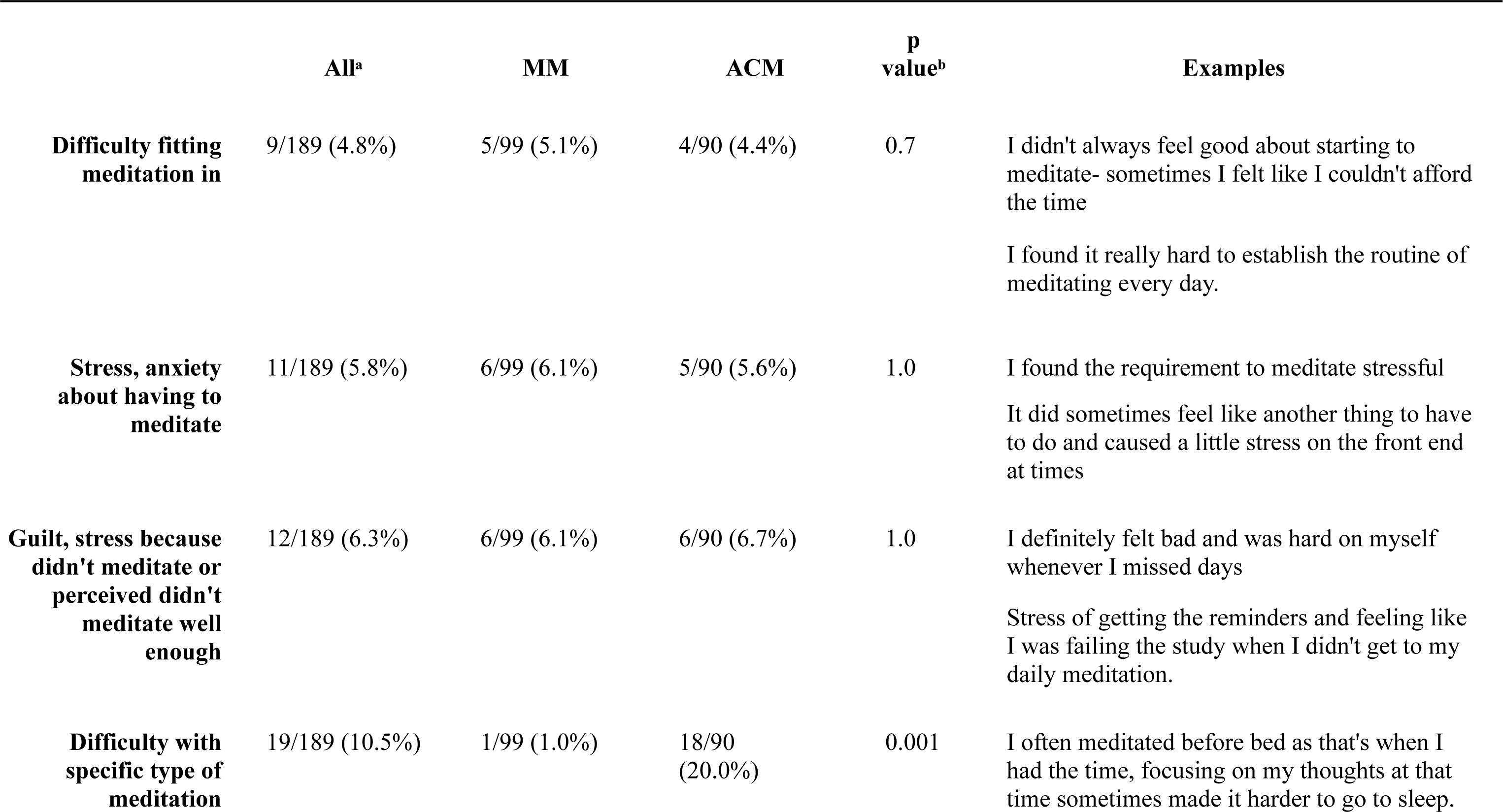
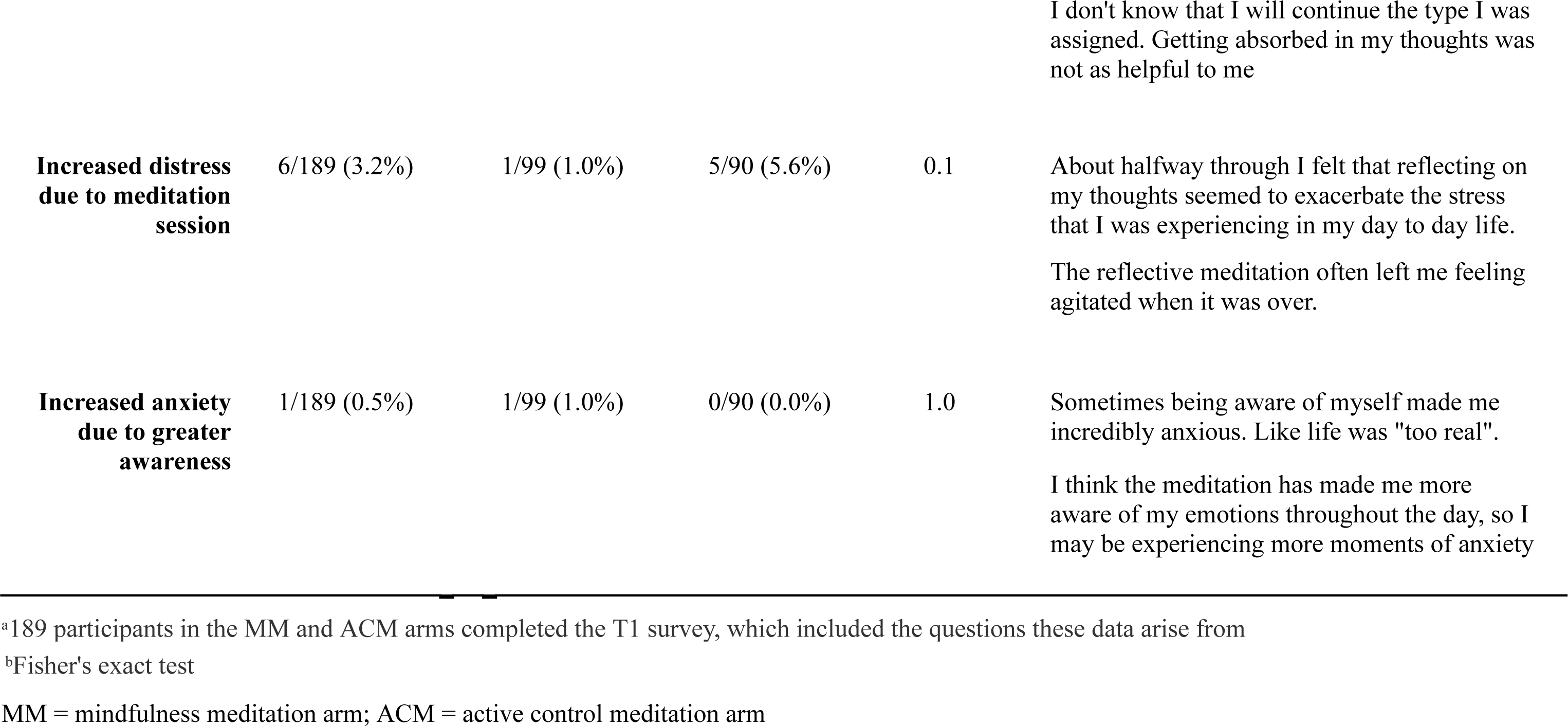
Participant-reported harms and negative effects.

|  | All<sup>a</sup> | MM | ACM | p<br>value<sup>b</sup> | Examples |
| --- | --- | --- | --- | --- | --- |
| **Difficulty fitting meditation in** | 9/189 (4.8%) | 5/99 (5.1%) | 4/90 (4.4%) | 0.7 | I didn't always feel good about starting to meditate- sometimes I felt like I couldn't afford the time<br>I found it really hard to establish the routine of meditating every day. |
| **Stress, anxiety about having to meditate** | 11/189 (5.8%) | 6/99 (6.1%) | 5/90 (5.6%) | 1.0 | I found the requirement to meditate stressful<br>It did sometimes feel like another thing to have to do and caused a little stress on the front end at times |
| **Guilt, stress because didn't meditate or perceived didn't meditate well enough** | 12/189 (6.3%) | 6/99 (6.1%) | 6/90 (6.7%) | 1.0 | I definitely felt bad and was hard on myself whenever I missed days<br>Stress of getting the reminders and feeling like I was failing the study when I didn't get to my daily meditation. |
| **Difficulty with specific type of meditation** | 19/189 (10.5%) | 1/99 (1.0%) | 18/90 (20.0%) | 0.001 | I often meditated before bed as that's when I had the time, focusing on my thoughts at that time sometimes made it harder to go to sleep. |

**Supplemental Table 7.** Five-Factor Mindfulness Questionnaire Subscale Scores by Timepoint and Study Arm.

| Arm | Subscale | T0 | T1 | p-value |
| --- | --- | --- | --- | --- |
| ACM | Observing | 23.9 | 27.4 | <0.0001 |
|  | Describing | 27.7 | 29.3 | 0.0007 |
|  | Acting with awareness | 23.6 | 26.1 | 0.0001 |
|  | Non-reactivity of inner experience | 19.8 | 22.7 | <0.0001 |
|  | Non-judging of inner experience | 25.3 | 28.9 | <0.0001 |
| NMC | Observing | 24.0 | 24.5 | 0.085 |
|  | Describing | 27.2 | 27.2 | 0.36 |
|  | Acting with awareness | 24.2 | 23.8 | 0.35 |
|  | Non-reactivity of inner experience | 19.5 | 20.1 | 0.026 |
|  | Non-judging of inner experience | 24.9 | 25.2 | 0.97 |
| MM | Observing | 23.9 | 27.2 | <0.0001 |
|  | Describing | 26.9 | 29.1 | 0.0001 |
|  | Acting with awareness | 23.5 | 25.3 | 0.017 |
|  | Non-reactivity of inner experience | 19.5 | 22.0 | <0.0001 |
|  | Non-judging of inner experience | 25.8 | 28.6 | 0.0003 |

Abbreviations: ACM, Active control Meditation; MM, Mindfulness Meditation; NMC, No-Meditation Control

**Supplemental Table 8.** Linear Regression Results for Intention-to-Treat Outcome Analyses for Burnout subscales.

| Outcome | Comparison | Term | Coefficient (β) | Standard Error | p-value |
| --- | --- | --- | --- | --- | --- |
| Work Exhaustion | MM vs ACM | Study Arm | -0.32 | 0.37 | 0.40 |
|  |  | T0 Work Exhaustion | 0.29 | 0.06 | < 0.001 |
|  |  | T0 Mindfulness | -0.01 | 0.05 | 0.88 |
|  | ACM vs NMC | Study Arm | 1.49 | 0.38 | < 0.001 |
|  |  | T0 Work Exhaustion | 0.46 | 0.06 | < 0.001 |
|  |  | T0 Mindfulness | 0.01 | 0.05 | 0.87 |
|  | MM vs NMC | Study Arm | -1.89 | 0.40 | < 0.001 |
|  |  | T0 Work Exhaustion | 0.43 | 0.06 | < 0.001 |
|  |  | T0 Mindfulness | -0.02 | 0.05 | 0.75 |
| Interpersonal Disengagement | MM vs ACM | Study Arm | -0.21 | 0.60 | 0.73 |
|  |  | T0 Interpersonal Disengagement | 0.26 | 0.07 | < 0.001 |
|  |  | T0 Mindfulness | 0.02 | 0.06 | 0.70 |
|  | ACM vs NMC | Study Arm | 1.84 | 0.56 | 0.002 |
|  |  | T0 Interpersonal Disengagement | 0.45 | 0.06 | < 0.001 |
|  |  | T0 Mindfulness | -0.06 | 0.06 | 0.28 |
|  | MM vs NMC | Study Arm | -2.07 | 0.56 | < 0.001 |
|  |  | T0 Interpersonal Disengagement | 0.36 | 0.06 | < 0.001 |
|  |  | T0 Mindfulness | -0.04 | 0.06 | 0.48 |

1. Abbreviations:
- MM: Mindfulness Meditation - ACM: Active Control Meditation - NMC: No-Meditation Control
2. Sample Size: N = 397

**Supplemental Table 9.**
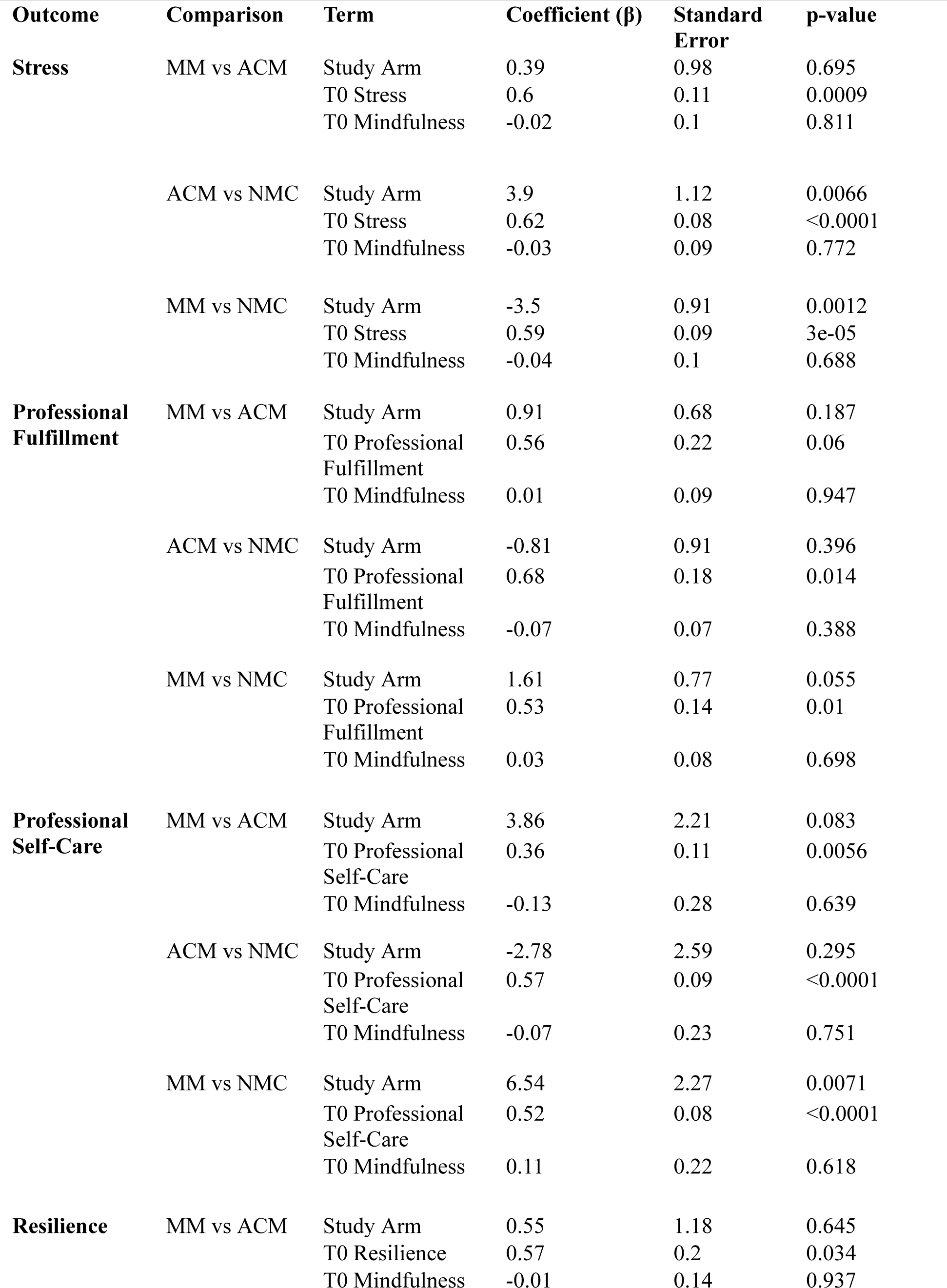
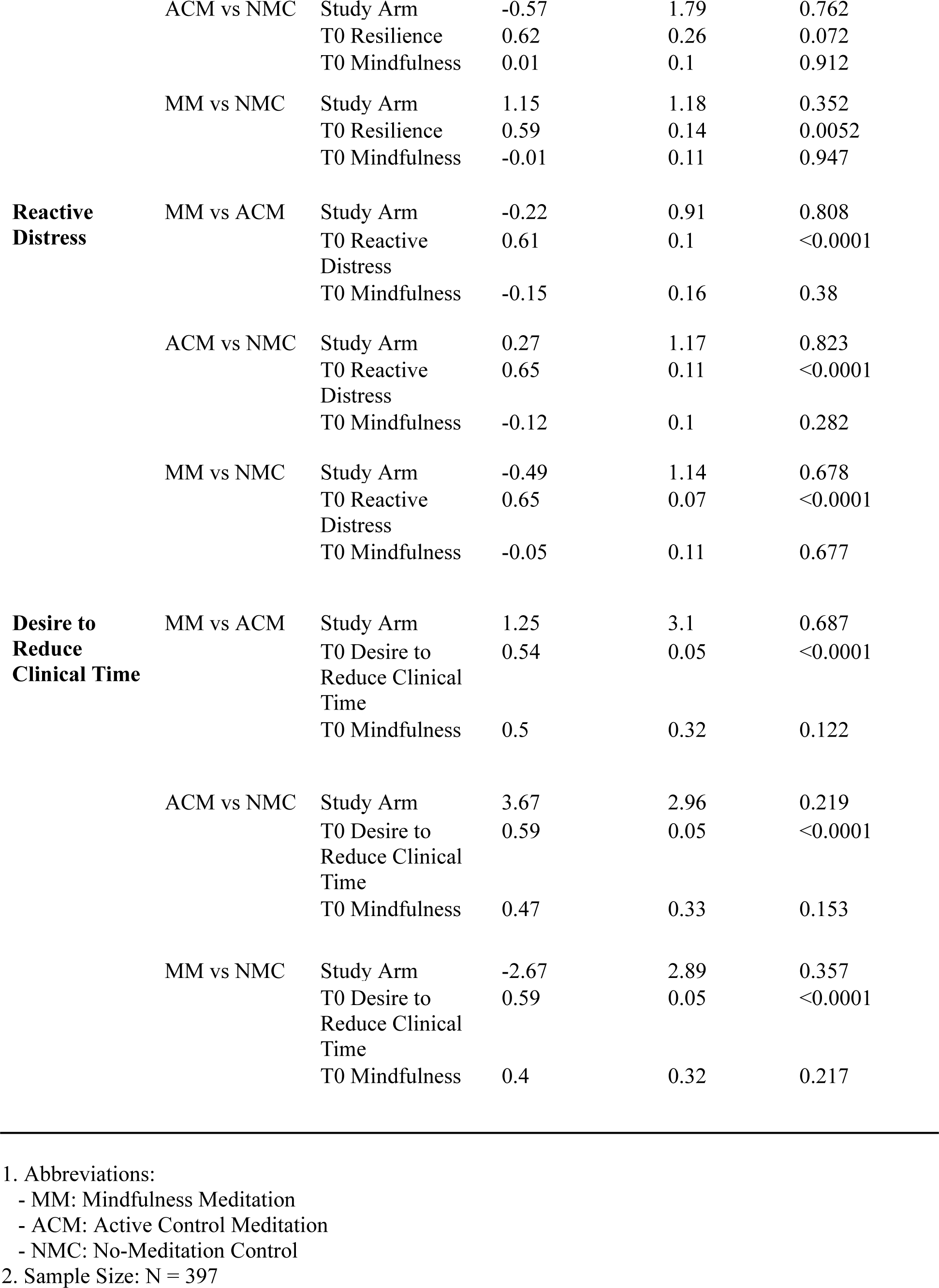
Linear Regression Results for Intention-to-Treat Outcome Analyses for Secondary Outcomes.

**Supplemental Table 10:** CONSORT 2025 checklist.

| Section/Topic | No | CONSORT 2025 checklist item description | Reported on page no. |
| --- | --- | --- | --- |
| **Title and abstract** |  |  |  |
| Title and structured abstract | 1a | Identification as a randomized trial | 1 |
|  | 1b | Structured summary of the trial design, methods, results, and conclusions | 2 |
| **Open science** |  |  |  |
| Trial registration | 2 | Name of trial registry, identifying number (with URL) and date of registration | 1 |
| Protocol and statistical analysis plan | 3 | Where the trial protocol and statistical analysis plan can be accessed | 1,7 |
| Data sharing | 4 | Where and how the individual de-identified participant data (including data dictionary), statistical code and any other materials can be accessed | 1 |
| Funding and conflicts of interest | 5a | Sources of funding and other support (eg, supply of drugs), and role of funders in the design, conduct, analysis and reporting of the trial | 1-2 |
|  | 5b | Financial and other conflicts of interest of the manuscript authors | 1 |
| **Introduction** |  |  |  |
| Background and rationale | 6 | Scientific background and rationale | 3 |
| Objectives | 7 | Specific objectives related to benefits and harms | 3 |
| **Methods** |  |  |  |
| Patient and public involvement | 8 | Details of patient or public involvement in the design, conduct and reporting of the trial | 7 |
| Trial design | 9 | Description of trial design including type of trial (eg, parallel group, crossover), allocation ratio, and framework (eg, superiority, equivalence, non-inferiority, exploratory) | 3 |
| Changes to trial protocol | 10 | Important changes to the trial after it commenced including any outcomes or analyses that were not prespecified, with reason | 4 |
| Trial setting | 11 | Settings (eg, community, hospital) and locations (eg, countries, sites) where the trial was conducted | 4 |
| Eligibility criteria | 12a | Eligibility criteria for participants | 5 |
|  | 12b | If applicable, eligibility criteria for sites and for individuals delivering the interventions (eg, surgeons, physiotherapists) | N/A |
| Intervention and comparator | 13 | Intervention and comparator with sufficient details to allow replication. If relevant, where additional materials describing the intervention and comparator (eg, intervention manual) can be accessed | 4 |
| Outcomes | 14 | Prespecified primary and secondary outcomes, including the specific measurement variable (eg, systolic blood pressure), analysis metric (eg, change from baseline, final value, time to event), method of aggregation (eg, median, proportion), and time point for each outcome | 6 |
| Harms | 15 | How harms were defined and assessed (eg, systematically, non-systematically) | 7 |
| Sample size | 16a | How sample size was determined, including all assumptions supporting the sample size calculation | 4 |
|  | 16b | Explanation of any interim analyses and stopping guidelines | N/A |
| **randomization:** |  |  |  |
| Sequence generation | 17a | Who generated the random allocation sequence and the method used | 5 |
|  | 17b | Type of randomization and details of any restriction (eg, stratification, blocking and block size) | 5 |
| Allocation concealment mechanism | 18 | Mechanism used to implement the random allocation sequence (eg, central computer/telephone; sequentially numbered, opaque, sealed containers), describing any steps to conceal the sequence until interventions were assigned | 5 |
| Implementation | 19 | Whether the personnel who enrolled and those who assigned participants to the interventions had access to the random allocation sequence | 5 |
| Blinding | 20a | Who was blinded after assignment to interventions (eg, participants, care providers, outcome assessors, data analysts) | 3-4 |
|  | 20b | If blinded, how blinding was achieved and description of the similarity of interventions | 4 |
| Statistical methods | 21a | Statistical methods used to compare groups for primary and secondary outcomes, including harms | 7 |
|  | 21b | Definition of who is included in each analysis (eg, all randomized participants), and in which group | 7 |
|  | 21c | How missing data were handled in the analysis | 7 |
|  | 21d | Methods for any additional analyses (eg, subgroup and sensitivity analyses), distinguishing prespecified from post hoc | 7 |
| **Results** |  |  |  |
| Participant flow, including flow diagram | 22a | For each group, the numbers of participants who were randomly assigned, received intended intervention, and were analysed for the primary outcome | 14 |
|  | 22b | For each group, losses and exclusions after randomization, together with reasons | 14 |
| Recruitment | 23a | Dates defining the periods of recruitment and follow-up for outcomes of benefits and harms | 5, 7 |
|  | 23b | If relevant, why the trial ended or was stopped | N/A |
| Intervention and comparator delivery | 24a | Intervention and comparator as they were actually administered (eg, where appropriate, who delivered the intervention/comparator, how participants adhered, whether they were delivered as intended (fidelity)) | 7-8 |
|  | 24b | Concomitant care received during the trial for each group | 8 |
| Baseline data | 25 | A table showing baseline demographic and clinical characteristics for each group | 15 |
| Numbers analysed, outcomes and estimation | 26 | For each primary and secondary outcome, by group:<br>• the number of participants included in the analysis<br>• the number of participants with available data at the outcome time point<br>• result for each group, and the estimated effect size and its precision (such as 95% confidence interval)<br>• for binary outcomes, presentation of both absolute and relative effect size | 8-9, 16 |
| Harms | 27 | All harms or unintended events in each group | 8, 24-25 |
| Ancillary analyses | 28 | Any other analyses performed, including subgroup and sensitivity analyses, distinguishing pre-specified from post hoc | 7-9 |
| **Discussion** |  |  |  |
| Interpretation | 29 | Interpretation consistent with results, balancing benefits and harms, and considering other relevant evidence | 9-12 |
| Limitations | 30 | Trial limitations, addressing sources of potential bias, imprecision, generalisability, and, if relevant, multiplicity of analyses | 12 |

